# Two-Person Psychopathology: Linguistic Style Matching Marks Disorganized Self-Referential Narrative in Psychosis

**DOI:** 10.64898/2026.08.24.26361227

**Authors:** Fiona Meister, Alban Voppel, Paulina Dzialoszynski, Lena Palaniyappan

**Author notes:** Corresponding Author: Dr. Lena Palaniyappan.

## Abstract

**Introduction:** Disturbed interpersonal attunement is a core but poorly operationalised feature of the psychopathology of schizophrenia. Language Style Matching (LSM), the largely unconscious alignment of two speakers’ function words during ordinary conversation, offers an observable, dialogue-derived index of this dyadic attunement. An open question is where altered alignment mark who a patient (a stable trait of inner experience) is or how they are (a fluctuating state that shifts with symptom severity)?

**Objective:** To characterise LSM over 12 months in early psychosis relative to controls, and to test, at both between- and within-person levels, whether alignment covaries with core psychopathological dimensions across self-referential (autobiographical) and externally directed discourse.

**Methods:** First-episode and recent-onset patients (n = 109) and controls (n = 60) completed semi- structured interviews at baseline and 12 months. LSM was computed per context and modelled with linear mixed-effects; a Mundlak decomposition partitioned the LSM–symptom association into between- and within-person components.

**Results:** LSM is not a fixed trait: groups were indistinguishable at baseline but diverged by 12 months (Group × Timepoint β = −0.018, p = .029), and the deficit was specific to autobiographical speech.

Within individuals, autobiographical alignment tightened as formal thought disorder rose above a patient’s own average and as negative symptoms worsened, independent of antipsychotic dose.

**Conclusion:** Patients aligned less than controls when speaking about themselves, yet aligned more as symptoms deteriorated — a shift from self-generated toward partner-scaffolded speech when self- organisation fails. LSM indexes disordered self-anchoring and interpersonal attunement in the negative–disorganized dimension of psychosis.

## Introduction

The capacity to align with a conversation partner’s linguistic choices is a fundamental feature of natural dialogue. Disruption of this alignment is an observable sign of communicative pathology that emerges in routine clinical encounters without requiring deliberate elicitation from the patient. Both excessive and deficient alignment carry communicative costs. Those abnormalities are particularly sailiant in certain clinical conditions such as schizophrenia where pragmatic, social-cognitive, or executive dimensions of language are disrupted [1–3]. Speaker alignment usually operates below awareness, especially at the level of function words which provide the grammatical scaffolding of an utterance. Language Style Matching (LSM) [4] has gained popularity to quantify this type of alignment. It aims to capture a dimension of syntactic-pragmatic coordination through which speaking partners construct shared common ground during conversation [5]. LSM can easily be derived from transcribed speech samples, making it a low-cost index of real-time interpersonal alignment and thus a prime candidate as an observable sign of communicative disruption in psychotic disorders.

Schizophrenia-spectrum disorders disrupt language through disorganized (e.g., formal thought disorder (FTD)) and impoverished thinking (e.g. reduced communicative initiative and negative symptoms) [6] which can both impair interpersonal alignment during interactions. However, previous studies concerned with speakers alignment yielded conflicting results: Reduced syntactic alignment between patients and interviewers has been observed, whereas no such reduction has been observed for lexical content-word alignment [7]. These findings suggest that alignment deficits are specific to structural-syntactic organization rather than propositional/semantic content. However, studies concerned with LSM specifically reported equally conflicting results: [8] recorded naturalistic everyday conversations and found modestly reduced LSM in established schizophrenia relative to healthy controls (4% reduction). This difference remained robust after comparing personal/emotionally laden conversation involving self-disclosure (e.g., “I feel upset today”) vs non- personal conversations (e.g., “How’s the weather”) where LSM was increased in the former compared to the latter. Another study [9] examined LSM during a structured role-play task (an interaction centered on external referents) in a first-episode psychosis sample and found no group difference, where individual variation in LSM was attributed to social functioning rather than diagnosis.

These divergent findings may not be contradictory but rather result from LSM being shaped by processes that operate at different levels of variation and present differently depending on discourse context: At the between-person level, negative symptoms and impoverished speech may reduce active engagement with a conversational partner’s speech, thus yielding lower LSM scores. At the within-person level, individual function-word production has been linked to symptom severity in first-episode schizophrenia [10], particularly to formal thought disorder (FTD) [11]. When FTD is pronounced, self-organized language structure may be disrupted [12], leaving speakers more reliant on the interviewer’s linguistic scaffold, thus yielding higher LSM scores. These two processes act at different levels of variation and are not mutually exclusive. Both would potentially be more apparent in autobiographical narratives, which require self-anchored speech production, than in externally directed discourse.The two-level account above implies that LSM’s clinical relevance may reside as much in within-person fluctuations as in between-group differences. However, such level of variation could not be detected by previous studies due to their cross-sectional between-person designs. In a cohort of first-episode and recent-onset patients followed over 12 months, the present study therefore combines longitudinal follow up, task-context comparison, and within-person analysis to address the following questions:

1. Whether patients show lower LSM than controls at baseline, and whether any group difference emerges or changes over a 12-month follow-up.
2. Whether any such deficit is context-specific, differentiating autobiographical (self-referent) from externally directed (other-referent) discourse.
3. Whether LSM tracks illness state as a within-person marker:

a. fluctuating with clinical state rather than reflecting stable individual differences, and
b. covarying with specific clinical dimensions beyond overall positive-symptom change.

## Methods

### Participants

English-speaking participants were recruited for the DISCOURSE in psychosis study [13] through the Prevention and Early Intervention Program for Psychoses (PEPP) at the London Health Sciences Centre, a defined catchment area program serving first-episode and recent-onset psychosis in Southwestern Ontario, Canada. Eligibility for the patient group required a diagnosis of psychosis established by a psychiatrist as per the Diagnostic and Statistical Manual of Mental Disorders (DSM) 5th edition criterion [14]. Participants with psychosis secondary to substance use disorder, an active substance dependence in the past year, or a neurological disorder affecting speech output were excluded. Healthy controls were recruited from the same geographic area through community advertising. Those with a personal or first-degree family history of psychosis were excluded.

At baseline, n = 109 patients and n = 60 healthy controls were enrolled (total n = 169). The two groups did not differ significantly in age or gender distribution. Usable transcripts were obtained for n = 99 patients and n = 57 healthy controls at baseline and for n = 70 patients and n = 45 healthy controls at 12 months (Figure S1). Analyses differed in their data requirements (baseline-only versus paired timepoints; transcript versus clinical completeness), so smaller numbers were available for some subanalyses. The relevant sample size is specified for each analysis. Retained and attrited participants did not differ in clinical characteristics (Supplement S1; Table S1). Written informed consent was obtained from all participants, and all procedures were approved by the Research Ethics Board at Western University (London, Ontario, Canada).

### Clinical Assessments

Symptom severity was assessed using the Positive and Negative Syndrome Scale (PANSS) [15], with a 6-item version focused on positive (P1: delusions; P2: Conceptual Disorganization; and P3: Hallucinatory Behavior) and negative symptoms (N1: Blunted Affect; N4: Passive/Apathetic Social Withdrawal; and N6: Lack of Spontaneity and Flow of Conversation) consisting of core features of remission and relapse [16]. FTD was rated using the Thought and Language Index (TLI) [17], which yields separate subscales for disorganization (Looseness, Peculiar Logic, etc.) and impoverishment of thinking (Poverty of Content, Preservation of Ideas, etc.). Global social and occupational functioning was rated using the Social and Occupational Functioning Assessment Scale (SOFAS) [18].

Antipsychotic medication load was quantified at each assessment using WHO-standardized Defined Daily Doses (DDD) [19], permitting cross-antipsychotic dose comparison. Finally, all ratings were completed at baseline and repeated at 12-month follow-up for patients

### Speech Data Collection

Trained research staff conducted interviews following the DISCOURSE semi-structured protocol [13]. Each interview comprised six sequential tasks designed to elicit speech: three self-referent autobiographical tasks (free speech, health questions, dream recall) and three other-referent externally structured (picture description, cartoon board, story recall). Prompts were delivered according to pre-established guidance. The full interview lasted 15–35 minutes and was repeated at 12-month follow-up. Audio was recorded using a SONY Stereo Digital Voice Recorder with Built-in USB, 4 GB (ICDPX470) placed centrally within 4 feet of both speakers. Transcripts were generated automatically in CHAT format using Batchalign [20] and verified manually against original recordings. Verified transcripts with diarized speaker labels are stored at talkbank.org/access/English/Palaniyappan/Discourse-UWO [13]. The full corpus comprised n=271 transcripts.

### LSM Computation

LSM was computed in Python 3.12.11 using Niederhoffer and Pennebaker’s [4] method for nine function-word categoriespersonal pronouns, impersonal pronouns, articles, numbers, auxiliary verbs, conjunctions, prepositions, negations, quantifiers). Proportional usage for each of the nine function words was determined separately for each interviewer/patient and interviewer/healthy control pair, using the following formula: LSM = 1 − |pct₁ − pct₂| / (pct₁ + pct₂ + 0.0001). LSM scores were then averaged across all nine function-word categories, yielding a single overall LSM score per interviewer pair, expressing whether interviewer and participant speech had comparable distributions of function words. Then within each discourse context (i.e., autobiographical vs. external tasks) all turns were concatenated before applying the formula, yielding one pooled LSM score per context, per session. All transcripts met a minimum 50-word threshold per speaker and per context. Examples of low-/high LSM scores are presented in Supplement S2, Table S2.

### Statistical Analysis

#### Primary Group Comparisons

Group differences in overall LSM were examined with linear mixed-effects modelling (LMER; random intercept per participant; fixed effects: group, timepoint, group × timepoint interaction; REML estimation Between-group comparisons at each timepoint were performed using independent samples t-tests. Within-group Baseline to 12-month change was computed through paired t-tests. Effect sizes are reported as Cohen’s d. LSM stability across the 12- month interval was estimated as the Pearson correlation between baseline and 12-month scores in the paired sample (Baseline + Follow-up LSM scores available, n=113). Paired clinical change from baseline to 12 months was examined in patients with complete paired clinical data using paired t-tests for each clinical variable (PANSS positive, PANSS negative, TLI impoverishment, TLI disorganization, and SOFAS), with Cohen’s d to measure effect size.

#### Context specificity

To test whether the group difference varied by discourse context across both timepoints (Group × Context interaction), a Generalized Estimating Equations (GEE) model added context (autobiographical vs. external) as a fixed effect, with baseline overall LSM as a covariate. Separate LMER models at baseline and 12 months then identified the timepoint at which the pattern emerged. The 12-month effect was confirmed in a GEE restricted to 12-month observations with task-matched baseline LSM as a covariate. Within each context, the group main effect was estimated from a baseline-adjusted LMER, while effect sizes were quantified as conventional independent-groups Cohen’s d on observed LSM pooled across timepoints. As a sensitivity check, verbal output (participant token count) was added as a covariate in a separate LMER to confirm the pattern was not driven by differences in speech quantity (Table S3).

#### Clinical state association

To test whether LSM covaries with clinical state within individuals, we reduced the five clinical variables to orthogonal (independant) dimensions by PCA (in the 90 patients with complete baseline ratings; see Participants) and modelled their longitudinal association with LSM. Sampling adequacy was confirmed by the Kaiser–Meyer–Olkin criterion and Bartlett’s test of sphericity; components were retained by the Kaiser criterion (eigenvalue > 1) and interpreted at absolute loadings ≥ 0.40 [[21,22]] (loadings in Table S4). Baseline loadings were applied to standardized 12-month values to project scores forward, so both timepoints contributed to each trajectory. Longitudinal associations were tested by LMER (random intercept per participant) with a Mundlak decomposition [23], partitioning each component’s association with LSM into a between-person term (person-mean, capturing stable individual differences) and a within-person term (deviation from person-mean, indexing state fluctuation around each patient’s own mean). Models used all available observations: The between-person term drew on the full sample (N = 90) and the within-person term on the 55 patients assessed at both timepoints (54 for external). If the within-person term remained significant with the between-person term included in the model, this would indicate that LSM covaries with clinical state change within individuals, rather than merely reflecting stable differences between participants.

#### LSM change and clinical trajectory

To test whether longitudinal change in LSM relates to a specific clinical course, we correlated LSM variations with clinical change in the 56 patients with complete paired clinical data. Orthogonal change dimensions were identified by PCA on the five within-patient change scores (12-month minus baseline) for PANSS positive, PANSS negative, TLI impoverishment, TLI disorganization, and SOFAS. Each clinical scores were z-scored and SOFAS sign were inverted so that higher values indexed worse functioning, aligning its direction with the four symptom scores.

Thus, components reflect the patterning of change across domains rather than absolute magnitude. Two components met the Kaiser criterion (eigenvalue > 1) and a third was retained on theoretical grounds, consistent with prior work distinguishing negative and disorganization symptom dimensions [24]. All three components were interpreted at |loading| ≥ 0.40 (Tables S5).

LSM change (ΔLSM = LSM_12m − LSM_BL) was corrected for regression to the mean by residualising ΔLSM on baseline LSM, separately per context (ΔLSM_adj; Figure S2), and correlated with change-PC scores (Pearson r; n = 55, as one patient lacked a usable LSM change score). This correlation was supported by performing two more analyses: (i) a GEE with a timepoint × PC interaction (exchangeable correlation, robust SE) as a confirmatory longitudinal test (Tables S6); (ii) a specificity check correlating each change-PC with change in antipsychotic load (ΔDDD, Table S7).

All significance thresholds were set at α = 0.05. Analyses were performed in Python 3.12.11 using statsmodels 0.14.6 (mixed-effects and GEE models), scikit-learn 1.7.2 (PCA), scipy 1.15.3 (correlations and group comparisons), and factor_analyzer 0.5.1 (KMO and Bartlett sampling-adequacy tests).

## Results

Sample characteristics: Descriptive statistics are reported in Table 1. Patients and healthy controls did not differ significantly in age (t = 0.29, p = .774) or gender distribution (χ² = 2.53, p = .112). As expected, groups differed significantly on all clinical measures. Patients showed markedly lower functional status (t = −13.70, p < .001) and greater thought disorder on the TLI impoverishment subscale (t = 4.30, p < .001). TLI disorganization did not differ significantly between groups.(t = 1.75, p = .082).

### Question 1 - Group differences in LSM and temporal emergence

As displayed in Figure 1, LMER of overall LSM (pooling autobiographical and external contexts) revealed a significant Group × Timepoint interaction (β = −0.018, SÈ= 0.008, p = .029), with however no significant Group (β = −0.016, p = .673) or Timepoint (β = +0.004, p = .509) effect. More specifically, overall LSM did not differ between groups at baseline (HC M = 0.850 ± 0.033, PT M = 0.843 ± 0.034; d = 0.20, p = .228) but was significantly lower in patients at 12 months (HC M = 0.854 ± 0.034, PT M = 0.830 ± 0.039; d = 0.66, p < .001). Patients also showed a within-group change in overall LSM (paired subsample: Δ = −0.017 ± 0.006 SEM, paired t(67) = 2.88, p = .005, d = −0.35) but controls remained stable (Δ = +0.005, p = .458).

**Fig. 1.**
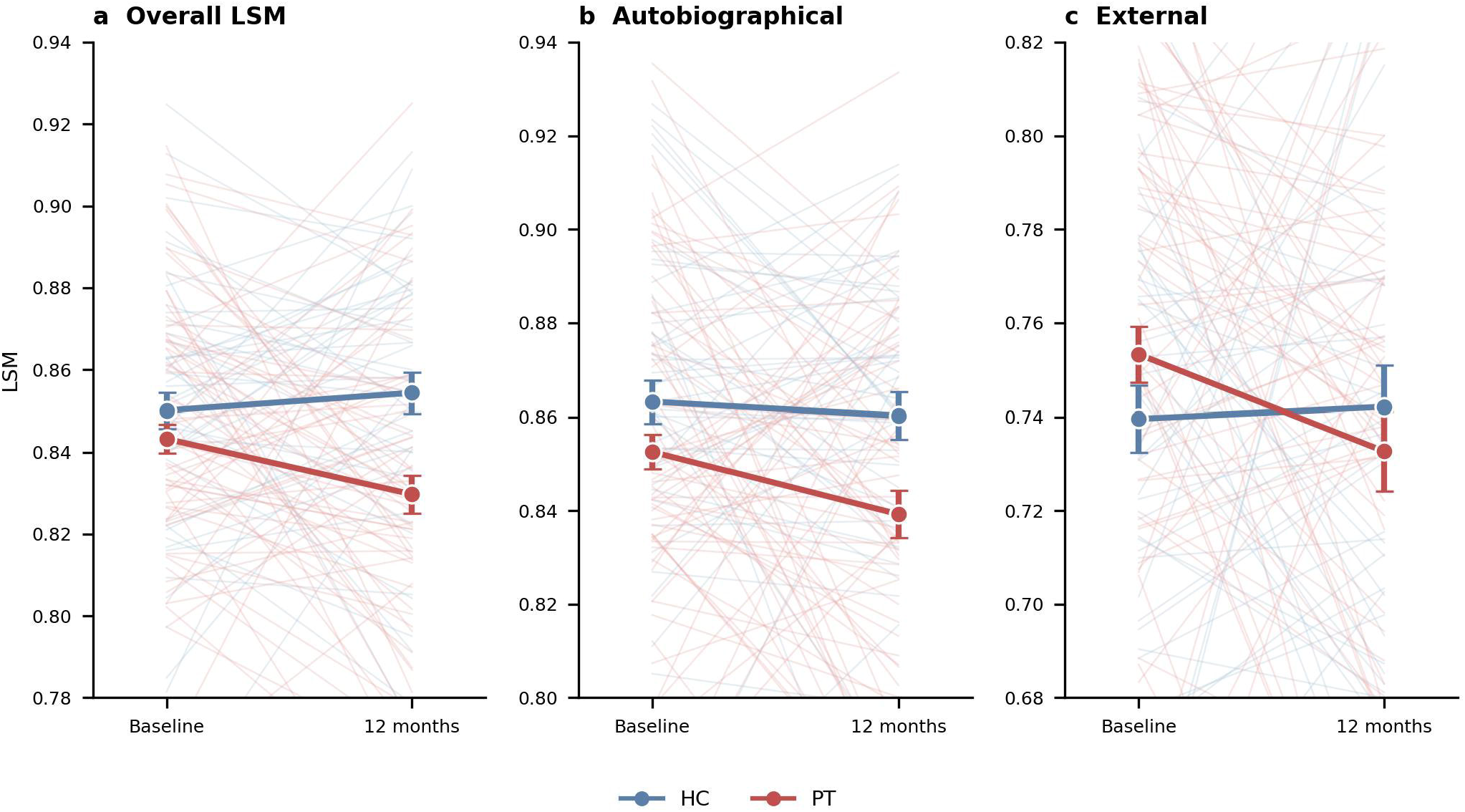
LSM trajectories from baseline to 12 months: Individual participant trajectories (faint lines) and group means (±SE, bold markers) are shown for overall LSM (left), autobiographical LSM (centre), and external LSM (right). Blue = healthy controls (HC); red = patients (PT). Note the y-axis range differs across panels (external LSM is lower in absolute terms); tick spacing is 0.02 throughout. HC, healthy controls; PT, patients. n = [HC / PT per timepoint].

### Question 2 - Context Specificity

As shown in Figure 2, a GEE extending the primary model (overall pooled scores) with discourse context (autobiographical vs. external) as a fixed effect revealed a significant Group × Context interaction across both timepoints (β = −0.019, SE = 0.009, p = .023). The group difference was significant for autobiographical (β = −0.016, p = .001; time-averaged d = 0.50) but not for external (β = +0.002, p = .857; d = −0.06) context. Those results suggest that the observed LSM differences between patients and controls was specific to autobiographical speech.

**Fig. 2.**
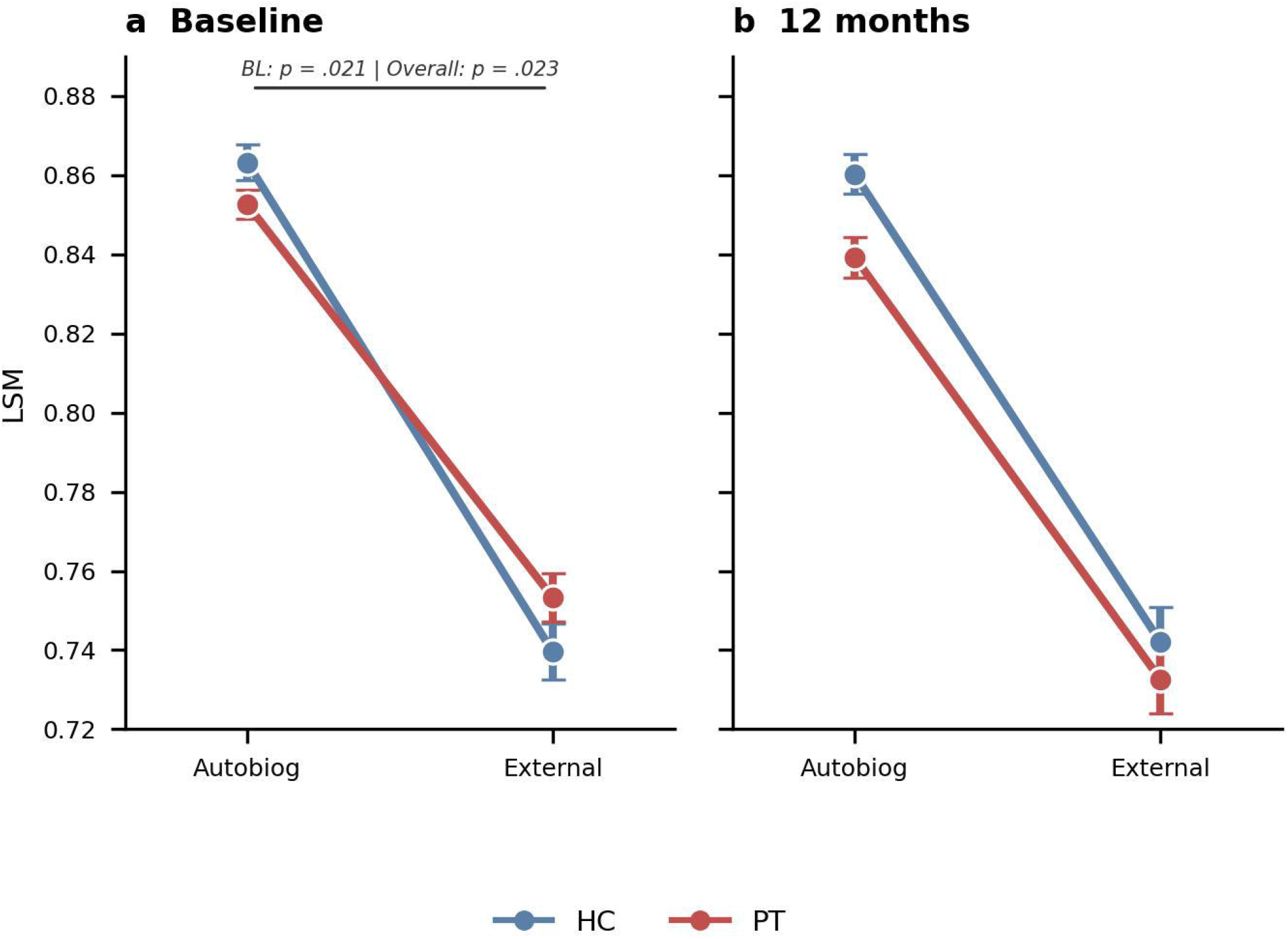
Extended LMER - Group × discourse context interaction: BL: baseline, HC: Healthy controls, PT: patients, LSM: language Style Matching. Group × discourse context interaction in linguistic style matching (LSM) at baseline (a) and 12-month follow-up (b). Points represent group means ± standard error.

To locate when this pattern emerged in time, we examined each timepoint separately (Figures 1–2). In autobiographical discourse, controls remained stable whereas patients’ LSM declined, so the group difference was present at baseline and widened by 12 months. In external discourse, patients were slightly higher than controls at baseline but slightly lower at 12 months, with controls again stable. As a result, the Group × Context interaction was significant at baseline (β = −0.024, SE = 0.011, p = .021): controls exceeded patients in autobiographical discourse, whereas patients slightly exceeded controls in external discourse, so the group difference reversed direction across contexts. By 12 months the interaction was non-significant (β = −0.012, SE = 0.013, p = .381), because patients now scored below controls in both contexts (the two differences shared the same direction and differed only in magnitude). Robustness checks adjusting for baseline LSM and verbal output are reported in Table S3.

### Question 3.a – LSM as a state marker

To determine whether LSM functions as a stable trait or a fluctuating state marker, we examined session-to-session stability across both timepoints.

Correlations of overall LSM (r = 0.15, p = .11, n = 113) and autobiographical LSM (r = 0.10, p = .27, n = 113) were not significant. Only external LSM reached statistical significance (r = 0.21, p = .029, n = 112), though the correlation was of similarly low magnitude to the two non-significant ones.. This indicates that single-session LSM scores are not stable trait estimates, and thus are possibly consistent sensitivity to state-.

To characterise clinical dimensions for the within-person analysis, PCA of the five baseline clinical variables (PANSS positive, PANSS negative, TLI impoverishment, TLI disorganization, and SOFAS) yielded two components by the Kaiser criterion (KMO = 0.644, Bartlett χ²(10) = 45.54, p < .001; Table S4), accounting for 59.8% of variance: PC1 (37.9%) indexed a severity-and-functioning axis (SOFAS +0.57, PANSS positive −0.53, PANSS negative −0.50), with higher scores indicating fewer symptoms and better global functioning. PC2 (21.9%) was dominated by TLI disorganization (+0.89), indexing formal thought disorder independently of overall severity (full component loadings: Table S4).

As illustrated in Figure 3, Mundlak LMER decomposed each clinical component into a between-person term (each patient’s average score across both timepoints) and a within-person term (deviation from that average at each assessment). Between-person variance was near-zero for overall and autobiographical LSM (random-intercept variance 0.00011–0.00012), indicating that stable individual differences in clinical profile did not drive alignment in these contexts. External discourse was the exception (random-intercept variance 0.00122, ICC = 0.32) and is discussed below. On the other hand, within-person changes in FTD (PC2) was associated with higher overall LSM (β = 0.019, SE = 0.005, d = 0.24, p = .002): when a patient’s thought disorder severity exceeded their own average, LSM increased. This effect was present in autobiographical discourse (β = 0.015, d = 0.19, p = .021), but absent for external discourse (β = 0.002, d = 0.01, p = .854). The severity-and-functioning axis (PC1) had no significant association with overall (p = .054), autobiographical (p = .120) or external context (p = .064) LSM.

**Fig. 3.**
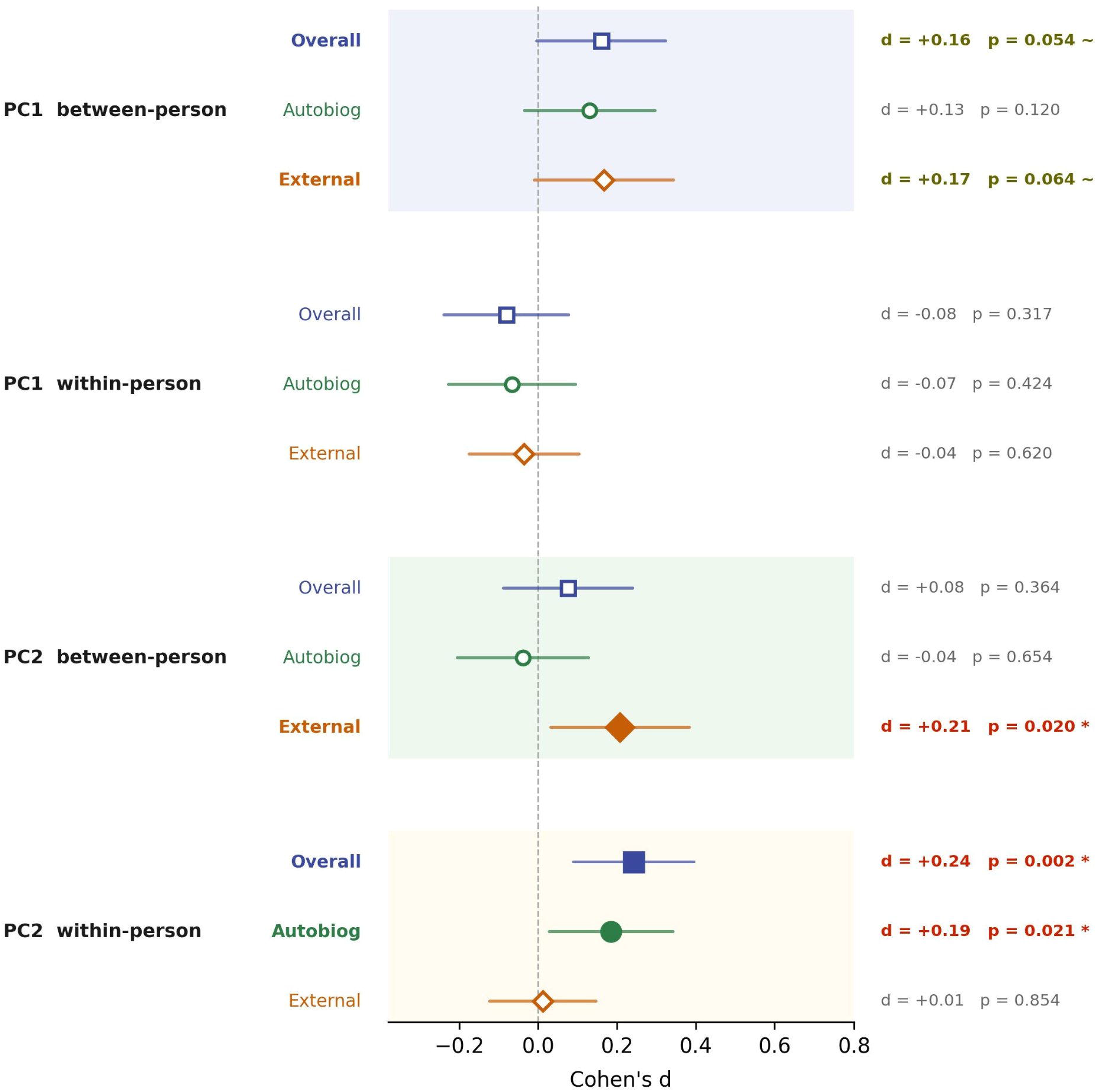
Forest plot of clinical predictors of linguistic style matching (LSM): PC = principal component; LSM = linguistic style matching; d = Cohen’s d. Mundlak LMER, patients only. Cohen’s d (signed) with 95% confidence intervals for between-person and within-person associations of baseline clinical PCA components (PC1: severity-and-functioning axis; PC2: formal thought disorder axis) with LSM across three discourse contexts. Marker shape indicates LSM context: square = overall, circle = autobiographical, diamond = external. Filled markers: p < .05; open markers: p ≥ .05; yellow-outlined markers: p < .10 (trend). Error bars = 95% confidence intervals.

The one exception to this within-person pattern was external discourse since the between-person component of formal thought disorder (PC2) was itself associated with LSM (β = 0.014, d = 0.21). Thus, external alignment was related to patients’ average disorganization but not their fluctuations (between-person significant, within-person null), whereas autobiographical alignment showed the opposite pattern (within-person significant, between-person null). This is consistent with external alignment being a stable, trait-like clinical profile and autobiographical alignment reflecting within-person state.

### Question 3.b – LSM and Clinical Changes

Because change scores are susceptible to regression to the mean, ΔLSM was corrected for this artefact prior to change-based analyses. Regression to the mean correction was applied after confirming that raw ΔLSM correlated with baseline LSM across all contexts (r = −0.48 to −0.67, all p < .001; Figure S2). After correction, residual correlations between baseline LSM and ΔLSM_adj were negligible (|r| < 0.001 in all contexts), which confirms the successful removal of the artefact.

Prior to the change-score PCA, patients showed modest clinical improvement over the 12-month follow-up, with significant reductions in PANSS positive (p = .004), PANSS negative (p = .014), and TLI disorganisation (p = .021). SOFAS showed a trend toward improvement (p = .055), whereas TLI impoverishment remained unchanged (p = .917). Those findings are consistent with clinical stabilization during follow-up despite possible individual exacerbations within the year.

Whereas we previously derived dimensions from baseline clinical levels, here we derived dimensions of clinical change: PCA of within-patient change scores yielded three components accounting for 76% of variance (Supplement S5; Table S5). The third component (eigenvalue = 0.919) was retained on theoretical grounds, consistent with prior literature distinguishing negative and disorganization symptom dimensions [24]. Change-PC1 (35%; Supplement S5; Table S5) loaded on PANSS-positive (+0.56; Figure 4), PANSS-negative (+0.50; Figure 4; Panel a), and SOFAS decline (+0.60; Figure 4; Panel a), indexing a general worsening axis on which all three clinical severity markers co-deteriorate.

**Fig. 4.**
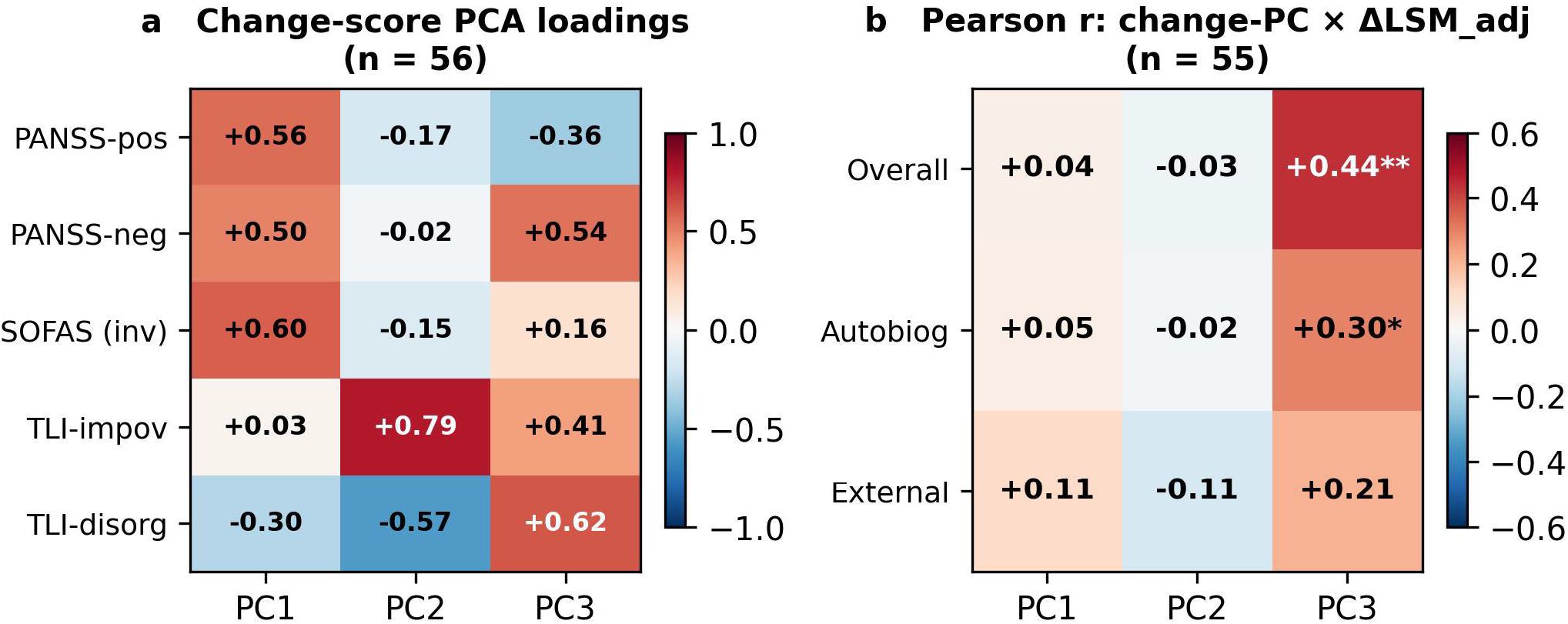
Change-score PCA loadings for the three retained components: PANSS-pos = Positive and Negative Syndrome Scale positive subscale; PANSS-neg = Positive and Negative Syndrome Scale negative subscale; SOFAS (inv) = Social and Occupational Functioning Assessment Scale, sign-inverted so that higher values indicate decline; TLI-impov = Thought and Language Index impoverishment subscale; TLI-disorg = Thought and Language Index disorganization subscale; PC = principal component; LSM = linguistic style matching;; RTM = regression to the mean; ΔLSM_adj = RTM- corrected LSM change score; r = Pearson correlation coefficient. ** p < .01; * p < .05. Panel a: Change-score PCA loadings for the three retained components (Change-PC1 35%, Change-PC2 23%, Change-PC3 18%). Positive values indicate relative worsening (compared to the general mean improvement) on that variable. Panel b: Pearson r between baseline corrected ΔLSM (rows) and each change-score PC (columns).

Change-PC2 (23%; Supplement S5; Table S5) was dominated by worsening TLI-impoverishment (+0.79; Figure 4) but improving TLI-disorganization (−0.57; Figure 4; Panel a), with negligible PANSS or SOFAS loadings, thus reflecting a shift in FTD rather than a change in severity per se (TLI-shift).

Finally, Change-PC3 (18%; Supplement S5; Table S5) loaded on TLI-disorganization (+0.62; Figure 4; Panel a), PANSS-negative (+0.54; Figure 4; Panel a), and TLI-impoverishment (+0.41; Figure 4; Panel a), indicating a combined worsening of disorganized and negative symptoms i.e., a negative- disorganized trajectory not associated with from the more overt worsening of positive symptoms.

To assess whether medication was associated with these clinical dynamics, change in antipsychotic load (ΔDDD = DDD at 12 months − DDD at baseline) was correlated with each change-PC (n = 46). The general-worsening Change-PC1 was associated with dose escalation (r = +0.33, p = .024), but the negative-disorganized Change-PC3 was not (r = −0.19, p = .20). 12-month antipsychotic load was also unrelated to 12-month LSM in any context (all |r| < 0.05, all p > .68; Table S7).

ΔLSM_adj was uncorrelated with Change-PC1 or Change-PC2 in any context (all |r| < 0.12, all p > .4). Change-PC3 was positively associated with overall (r = +0.44, p = .001) and autobiographical (r = +0.30, p = .028) ΔLSM_adj, but not external (r = +0.21, p = .131; Figure 4, Panel b). This pattern was confirmed in GEE analyses (Table S6).

## Discussion

In this study, we investigated the association between LSM and clinical trajectory in schizophrenia-spectrum disorders. Four main findings emerged. First, patients did not differ from healthy controls at baseline but developed significantly lower overall LSM over the subsequent 12 months. Second, this group difference was sensitive to speech task context, as it occurred only in autobiographical and not external discourse. Third, LSM exhibited minimal stability across repeated assessments and covaried with within-person fluctuations in FTD rather than stable trait-like differences between individuals. Finally, longitudinal increases in LSM were associated with worsening negative and disorganization symptoms, regardless of antipsychotic dose changes. By contrast, no relationship was found between general symptomatic worsening and LSM. These findings are consistent with LSM being a possible state-sensitive socio-behavioral marker rather than a fixed characteristic of schizophrenia-spectrum disorders.

The central result is that LSM moved in opposite directions at two levels of analysis: Between groups, patients aligned less than controls, specifically when speaking about themselves. On change within persons, the relationship reversed: as a patient’s FTD rose from their own average, and as negative and disorganization symptoms worsened over follow-up, alignment increased. These are complementary rather than conflicting results. We interpret the between-group deficit as reduced self-anchored linguistic engagement: On average, patients produce less of the self-referential language that autobiographical discourse demands. By contrast, the within-person increase reflects compensatory mechanisms: When the capacity to structure one’s own discourse is impaired, function-word use drifts toward the partner’s, thus raising LSM. LSM therefore does not rank speakers from worse to better alignment but rather is associated with the degree to which a speaker’s output is shaped by its partner rather than internally generated. This explains why LSM can be lower overall in patients than controls yet rise within a patient as their condition worsens.

These findings help reconcile two apparently conflicting prior reports. [9] found no group difference in LSM using a structured social role-play. Although their task differs from ours, it shares an externally imposed structure, matching our own finding that the group difference disappeared in external discourse. Their null result is therefore consistent with our results, since we found no group difference in externally scaffolded speech either. By contrast, [8] found a modest reduction in naturalistic LSM in established illness that weakened once symptom severity was taken into account. Our two-timepoint design indicates that this change could lie in the joint worsening of negative and disorganization symptoms, not in overall severity, a distinction their cross-sectional design could not capture. Taken together, what appeared to be conflicting results across studies in fact reflects the context- and dimension-specificity of the effect.

Two further results support this interpretation. First, the between- and within-person effects differed by context: external alignment was related to a patient’s average level of disorganization but not its fluctuations, whereas autobiographical alignment showed the opposite. This pattern of associations - can be used to map external discourse onto stable, trait-like clinical characteristics and autobiographical discourse onto within-person state, consistent with the trait-versus-state distinction seen throughout. Second, the longitudinal signal was specific to a clinical profile: LSM change was associated with the negative–disorganized trajectory but not general symptomatic worsening, and was independent of antipsychotic dose change, which instead accompanied the general-worsening dimension. Together, these findings suggest that LSM could capture a subtle dimension of disorganization and negative symptomatology that was not reflected in medication adjustments over the same interval.

Our study presents several limitations. Because LSM is a property of the dyad and interviewer speech was not controlled, we cannot exclude that some variation reflects interviewers adapting to patients rather than the reverse. The analytic sample was smaller than our total sample for some analyses, and the within-person and prospective estimates rest on the subset assessed at both timepoints (n = 55–56), which limits statistical power. The change component carrying the prospective signal (Change-PC3) was also retained below the conventional eigenvalue threshold and should be regarded as exploratory. Effect sizes were modest and not corrected for multiple comparisons.

Because low test–retest stability could reflect measurement noise, our state interpretation rests on the within-person clinical associations rather than on the instability itself. Finally, the data are single-site and semi-structured rather than fully naturalistic.

Taken together, LSM behaved as a state-sensitive marker rather than a fixed trait, tied to the disorganized and negative dimension of psychosis. Patients aligned less than controls when speaking about themselves, but aligned more as their symptoms worsened, interpreted as leaning on the interviewer’s language when their own became harder to organize. Because LSM can be measured automatically from ordinary conversation, with no specific self-report required, it could serve as a practical, low-burden marker of clinical state. Confirming its reliability in individual patients, and how much it depends on the conversation partner, will be important next steps in the field of dyadic (or two-person) psychopathology.

## Statements

### Statement of Ethics

#### Study approval statement

This study protocol was reviewed and approved by the Research Ethics Board at Western University, Ontario, Canada [Project ID: 119548].

#### Consent to participate statement

Written informed consent was obtained from all patients/participants prior to participation to the study. Research Ethics Board at Western University approved all study procedures. Helsinki statement

## Data Availability

Access to the dataset used in this article can be requested at: https://talkbank.org/psychosis/access/English/Palaniyappan/Discourse-UWO.html

## Acknowledgment

We thank the physicians, staff, and patients at the Prevention and Early Intervention Program for Psychoses at London Health Sciences Centre and Dr. Rohit Lodhi for clinical recruitment efforts. We thank the DISCOURSE consortium (https://discourseinpsychosis.org/) Steering Group for their assistance in developing the speech assessment protocol. We acknowledge Prof. Brian MacWhinney of Carnegie Mellon University for supporting the transcription and curation of this dataset and establishing the psychosis subsection of TalkBank to host DISCOURSE consortium data. We thank Chaimaa El-Mouslih and Dr. Michael Mackinley (London, Ontario) for assistance with transcript curation. Manuscript preparation was assisted by Claude (Anthropic, claude.ai; Sonnet 4.6), which was used for code review, statistical output verification, and editorial support. All analyses, interpretations, and final manuscript content were reviewed and approved by the authors who take full responsibility for the content.

## Funding Sources

### Author Contributions

**FM:** Data curation, writing - original draft preparation, review & editing. **AV:** Data Curation Supervision. PD: Data Collection. **LP:** Conceptualization, methodology, software, writing – Original draft preparation, review & editing, supervision, project administration.

## References

1. Agostoni G, Bambini V, Bechi M, et al. Communicative-pragmatic abilities mediate the relationship between cognition and daily functioning in schizophrenia. Neuropsychology. 2021;35(1):42–56. doi:10.1037/neu0000664

2. Gariup M, Bergström T, Saliger K, et al. Enhancing social cognition in psychosis – the potential role of open dialogue. Schizophrenia. 2025;11(1):84. doi:10.1038/s41537-025-00608-y

3. Meister F, Sellier Silva M, Melshin G, et al. Expressive pragmatic language in mood and psychotic disorders: a systematic review and meta-analysis. Schizophrenia. 2026;12(1):31. doi:10.1038/s41537-026-00733-2

4. Niederhoffer K, Pennebaker J. Linguistic Style Matching in Social Interaction. J Lang Soc Psychol. 2002:337–360. doi:10.1177/026192702237953

5. Pickering MJ, Garrod S. Toward a mechanistic psychology of dialogue. Behav Brain Sci. 2004;27(2):169–190; discussion 190-226. doi:10.1017/s0140525x04000056

6. Palaniyappan L, Homan P, Alonso-Sanchez MF. Language Network Dysfunction and Formal Thought Disorder in Schizophrenia. Schizophr Bull. 2022;49(2):486–497. doi:10.1093/schbul/sbac159

7. Sharpe V, Schoot L, Lewandowski KE, et al. We both say tomato: Intact lexical alignment in schizophrenia and bipolar disorder. Schizophr Res. 2022;243:138–146. doi:10.1016/j.schres.2022.02.032

8. Abel DB, Myers EJ, Whan BA, et al. Real-world conversations across the schizophrenia spectrum: Implementing passive audio sensing to examine linguistic style matching. J Psychopathol Clin Sci. 2025;134(8):872–881. doi:10.1037/abn0000998

9. Angers K, Kilicoglu MFV, Luck J, Zaccaria B, Vaughn KE, Moe AM. Conversationally Attuned: Links Between Interpersonal Linguistic Synchrony and Social Functioning Across the Early-Psychosis Spectrum. Psychiatry. Published online May 8, 2026:1–16. doi:10.1080/00332747.2026.2660514

10. Dalal TC, Park MTM, Silva AM, et al. Clinical psychopathology-based early relapse prediction model using speech and language in psychosis. Schizophr Res Cogn. 2025;43:100392. doi:10.1016/j.scog.2025.100392

11. Silva A, Limongi R, MacKinley M, Palaniyappan L. Small Words That Matter: Linguistic Style and Conceptual Disorganization in Untreated First-Episode Schizophrenia. Schizophr Bull Open. 2021;2(1):sgab010. doi:10.1093/schizbullopen/sgab010

12. Elleuch D, Chen Y, Luo Q, Palaniyappan L. Speaking of Yourself: A Meta-Analysis of 80 Years of Research on Pronoun Use in Schizophrenia. 2024.

13. Cho B, Balles E, Mackinley M, et al. The DISCOURSE in psychosis (London Ontario): A speech dataset to examine communication disturbances in early-stage psychosis. Data Brief. 2026;65:112517. doi:10.1016/j.dib.2026.112517

14. Association AP. The Diagnostic and Statistical Manual of Mental Disorders, Fifth Edition, Text Revision. Published online 2022. Accessed April 7, 2025. https://www.psychiatry.org:443/psychiatrists/practice/dsm

15. Kay SR, Fiszbein A, Opler LA. The positive and negative syndrome scale (PANSS) for schizophrenia. Schizophr Bull. 1987;13(2):261–276. doi:10.1093/schbul/13.2.261

16. Østergaard SD, Lemming OM, Mors O, Correll CU, Bech P. PANSS-6: a brief rating scale for the measurement of severity in schizophrenia. Acta Psychiatr Scand. 2016;133(6):436–444. doi:10.1111/acps.12526

17. Liddle PF, Ngan ETC, Caissie SL, et al. Thought and Language Index: an instrument for assessing thought and language in schizophrenia. Br J Psychiatry. 2002;181(4):326–330. doi:10.1192/bjp.181.4.326

18. Samara MT, Engel RR, Millier A, Kandenwein J, Toumi M, Leucht S. Equipercentile linking of scales measuring functioning and symptoms: examining the GAF, SOFAS, CGI-S, and PANSS. Eur Neuropsychopharmacol J Eur Coll Neuropsychopharmacol. 2014;24(11):1767–1772. doi:10.1016/j.euroneuro.2014.08.009

19. World Health Organization. The ATC/DDD Methodology. Accessed May 21, 2026. https://www.who.int/tools/atc-ddd-toolkit/methodology

20. Liu H, MacWhinney B, Fromm D, Lanzi A. Automation of Language Sample Analysis. J Speech Lang Hear Res JSLHR. 2023;66(7):2421–2433. doi:10.1044/2023_JSLHR-22-00642

21. Hair JF, Black WC, Babin BJ, Anderson RE. Multivariate Data Analysis. Eighth edition. Cengage; 2019.

22. Zwick WR, Velicer WF. Comparison of five rules for determining the number of components to retain. Psychol Bull. 1986;99(3):432–442. doi:10.1037/0033-2909.99.3.432

23. Mundlak Y. On the Pooling of Time Series and Cross Section Data. Econometrica. 1978;46(1):69–85. doi:10.2307/1913646

24. Liddle PF. The symptoms of chronic schizophrenia. A re-examination of the positive-negative dichotomy. Br J Psychiatry J Ment Sci. 1987;151:145–151. doi:10.1192/bjp.151.2.145

