## Supplementary Material for "Two-Person Psychopathology: Linguistic Style Matching Marks Disorganized Self-Referential Narrative in Psychosis"

#### **S1 Attrition/Retention**

Of 109 patients enrolled, 99 provided usable baseline transcripts (10 excluded for absent or unusable recordings). Of those 99, 68 returned, along with 2 more without baseline transcripts but returned at 12 months (total n=70; 71%). Of 60 healthy controls enrolled, 57 provided baseline transcripts and 45 (79%) returned at 12 months. Figure S1 illustrates participants attrition/retention between baseline and 12-month follow-ups. The groups were broadly comparable with no significant difference in clincial characteristics (see Table S1).

100% of subjects with usable transcripts at baseline had PANSS scores, while TLI was not available for 1 subject who did not complete picture description tasks. 8 individuals at baseline did not have sufficient information to score SOFAS. In total 90 patients at baseline had sufficient data to characterize clinical severity. Of 70 patients with 12-months data, 68 had both baseline and 12m transcripts; as 13 had no SOFAS scores, n=55 had sufficient data to characterize clinical changes over 1 year.

**S2 LSM Computation**

LSM score were computed using the following formula**:** LSM = 1 − |pct₁ − pct₂| / (pct₁ + pct₂ + 0.0001) where pct_1/2_ denote the proportional usage of each function-word category (as a percentage of total words) by the interviewer and participant respectively. The small constant (0.0001), as specified in the original LSM algorithm, prevents division by zero when neither speaker uses a given function-word category. Table S2 presents concrete examples of LSM calculations using transcripts from our sample, contrasting low- versus high-LSM, respectively.

**S3 Verbosity Control**

To examine whether group differences in LSM were attributable to differences in verbal output, a linear mixed-effects model (with a random intercept per participant) was used to test whether participant token count predicted LSM independent of timepoint. Token count was z-scored and timepoint was centred at 0. Models were run separately for each discourse context in patients only. As shown in Table S3, participant word output significantly predicted autobiographical LSM (β = 0.009, d = 0.23, p = .002) and overall LSM (β = 0.011, d = 0.29, p < .001), but not external LSM (p = .445). The context-specific group difference therefore cannot be explained by verbosity alone.

**S4 Baseline Clinical Principal Component Analysis**

A principal component analysis (PCA) was conducted on five z-scored baseline clinical variables in the 90 patients with complete data: PANSS positive, PANSS negative, TLI impoverishment, TLI disorganization, and SOFAS. Sampling adequacy was confirmed (KMO = 0.644; Bartlett's χ²(10) = 45.54, p < .001). Two components met the Kaiser criterion (eigenvalue > 1), together accounting for 59.8% of the variance (Table S4). BL-PC1 represented a general severity/functioning dimension, whereas BL-PC2 primarily reflected formal thought disorder. The baseline loading matrix was subsequently applied to standardized 12-month clinical values to compute projected follow-up component scores.

**S5 Change-score PCA**

To identify orthogonal dimensions of clinical change, PCA was performed on five standardized within-patient change scores (12-month − baseline). SOFAS was sign-inverted so that higher values reflected worsening. Sampling adequacy was acceptable (KMO = 0.589; Bartlett's χ²(10) = 22.14, p = .014). Two components met the Kaiser criterion, while a third component (eigenvalue = 0.919) was retained on theoretical grounds because it represented a clinically interpretable negative-disorganization trajectory. The resulting component structure is shown in Table S5.

**S6 Prospective associations between clinical change and LSM**

Prospective associations between clinical change dimensions and RTM-corrected LSM change were evaluated using two complementary analyses. Pearson correlations quantified the association between each change-PC score and ΔLSM_adj within each discourse context, whereas generalized estimating equations (GEE) provided a confirmatory longitudinal analysis using the timepoint × PC interaction. Results from both analyses are summarized together in Table S6. Across both approaches, only cPC3 (negative-disorganisation trajectory) showed consistent associations with LSM change.

**S7 Regression-to-the-Mean Correction**

LSM change scores (ΔLSM = LSM_12m − LSM_BL) correlated strongly with baseline LSM in both patients and healthy controls across all three contexts (r = −0.48 to −0.67, all p < .001; Figure S2), indicating a substantial regression-to-the-mean (RTM) artefact whereby participants with higher baseline LSM showed greater apparent decline and those with lower baseline LSM showed apparent improvement. To remove this artefact, RTM-corrected change scores (ΔLSM_adj) were computed as residuals from a linear regression of ΔLSM on baseline LSM, fitted separately for each group and linguistic context. After correction, residual correlations between baseline LSM and ΔLSM_adj were negligible (|r| < 0.001 in all contexts; Figure S2), confirming successful removal of the artefact. All prospective analyses involving LSM change use ΔLSM_adj rather than raw ΔLSM.

**S8 Medication Specificity**

To confirm that change-PC scores reflected clinically meaningful trajectories rather than medication adjustment effects, each component was correlated with changes in antipsychotic dose expressed as defined daily doses (ΔDDD = DDD_AP_12m − DDD_BL) in the 46 patients with complete PCA and medication data. Antipsychotic load did not change significantly over the follow-up period (mean ΔDDD = −0.107, SD = 0.739; paired t = 0.021, p = .984; Wilcoxon p = .919), and 12-month DDD was unrelated to 12-month LSM in any context (all |r| < 0.05, all p > .68). As shown in Table S7, only cPC1 (general improvement) correlated significantly with ΔDDD (r = +0.333, p = .024), indicating that clinicians increased antipsychotic dose in patients showing less clinical improvement - a pattern consistent with ineffective treatment or relapse as recognized by the treating clinicians. cPC2 and cPC3 were not associated with medication change (both p > .19), suggesting that the negative-disorganisation trajectory captured by cPC3 was clinically not salient with respect to pharmacological management.

**Figures**:

**Fig. S1: Sankey diagram of longitudinal data acquisition and attrition.** *BL: baseline, HC: Healthy controls, PT: patients, LSM: language Style Matching*

**Fig. S2:** **Scatterplots of raw LSM change (ΔLSM) as a function of baseline LSM**. *BL: baseline; HC: healthy controls; PT: patients; LSM: linguistic style matching; Δ: change score (12-month minus baseline); RTM: regression to the mean; β: unstandardized regression coefficient; r: Pearson correlation coefficient; p: p-value. ΔLSM are shown separately for healthy controls (top row, blue) and patients (bottom row, red), and across the three discourse contexts (autobiographical, external, overall).*
