## Supplementary material for "Two-Person Psychopathology: Linguistic Style Matching Marks Disorganized Self-Referential Narrative in Psychosis": Table S4

| **Component** | **Eigenvalue** | **% Variance** | **Cumulative %** | **PANSS Positive** | **PANSS Negative** | **SOFAS** | **TLI Impoverishment** | **TLI Disorganisation** |
| --- | --- | --- | --- | --- | --- | --- | --- | --- |
| **BL-PC1** | **1.916** | **37.9** | **37.9** | **-0.525** | **-0.502** | **+0.569** | -0.385 | +0.012 |
| **BL-PC2** | **1.107** | **21.9** | **59.8** | -0.028 | -0.348 | -0.196 | +0.232 | **+0.887** |

**Note**. Principal component analysis (PCA) was performed on five baseline clinical variables in 90 patients: PANSS positive, PANSS negative, TLI impoverishment, TLI disorganisation, and SOFAS. Sampling adequacy was acceptable (KMO = 0.644; Bartlett's χ²(10) = 45.54, p < .001). Two components met the Kaiser criterion (eigenvalue > 1) and were retained. Component loadings were extracted without rotation. Bold values indicate retained components and salient loadings (|loading| ≥ 0.40). BL-PC1 contrasts higher SOFAS (better functioning) with greater PANSS positive and negative symptom burden, reflecting a general severity/functioning dimension. BL-PC2 primarily loads on TLI disorganisation, reflecting a formal thought disorder dimension. The baseline loading matrix was subsequently applied to standardized 12-month clinical variables to compute projected follow-up component scores (n = 56); no separate PCA was performed at follow-up.

**Table S4. Baseline Clinical PCA**
