## Supplementary figures and images for "Two-Person Psychopathology: Linguistic Style Matching Marks Disorganized Self-Referential Narrative in Psychosis"

### Figure S1

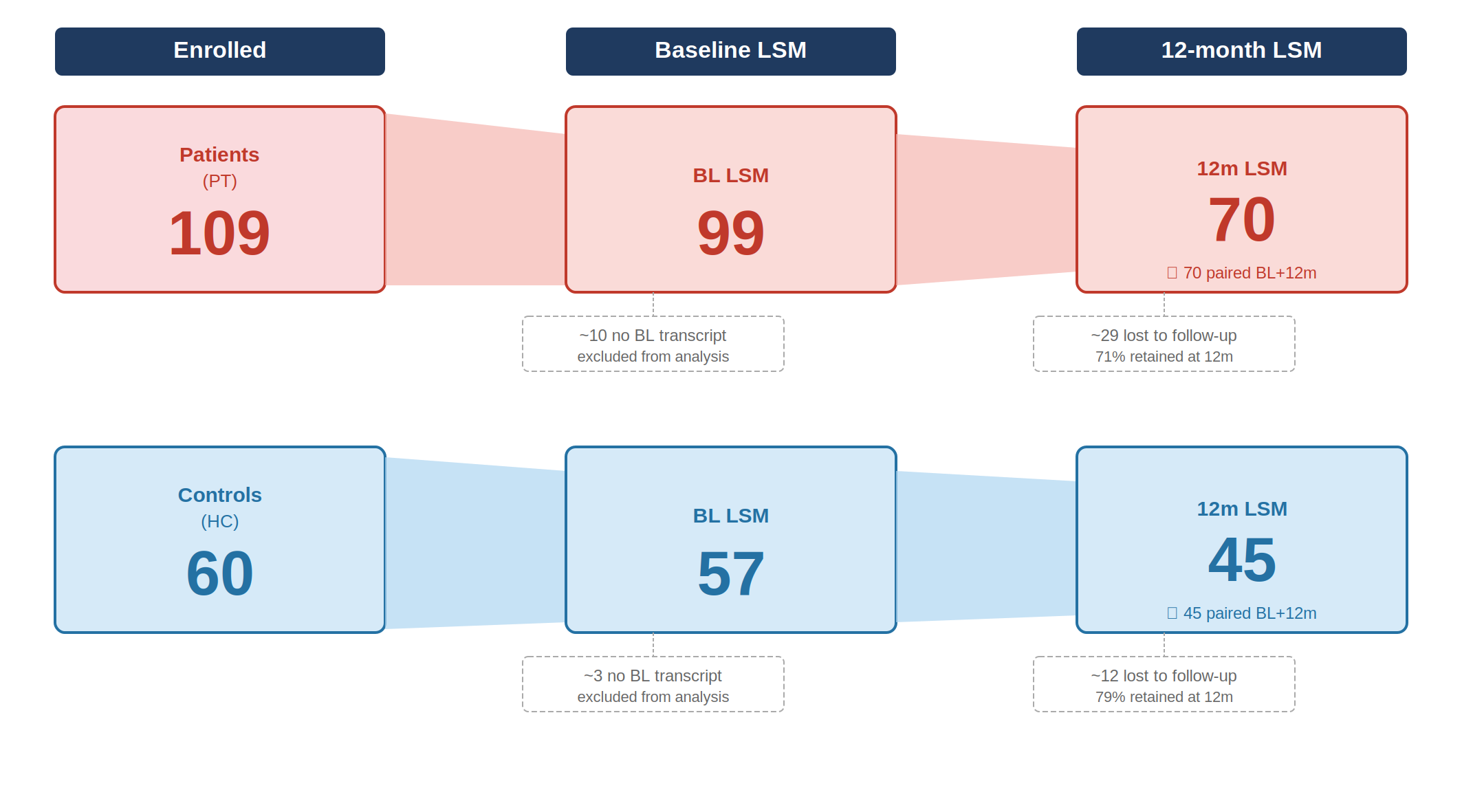

### Figure S2

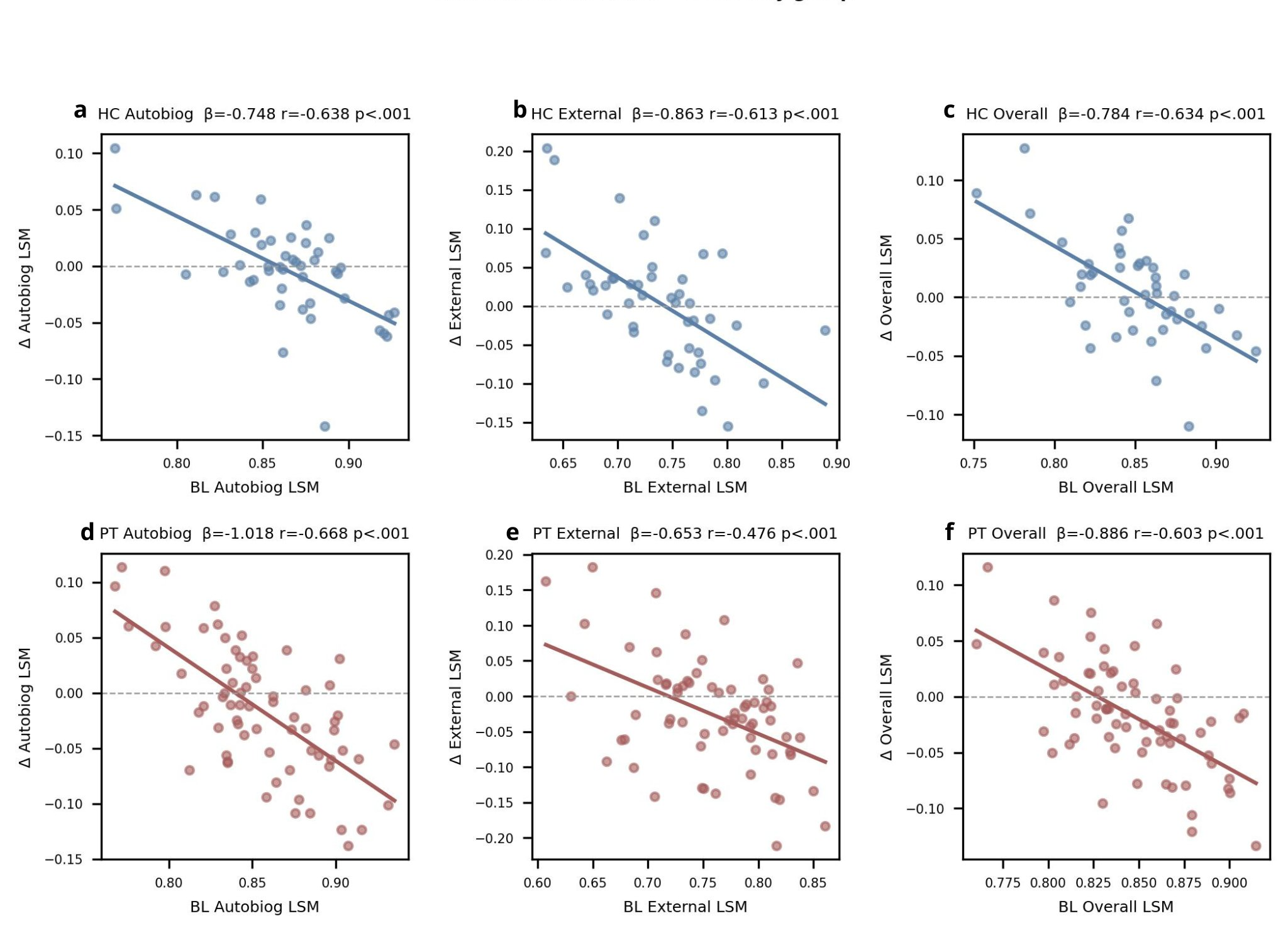
