## Supplementary material for "Two-Person Psychopathology: Linguistic Style Matching Marks Disorganized Self-Referential Narrative in Psychosis": Table S1

| **Variable** | **Retained (n=70)** | | **Attrited (n=39)** | | **t** | **p** | **d** | **Sig.** |
| --- | --- | --- | --- | --- | --- | --- | --- | --- |
|  | **M** | **SD** | **M** | **SD** |  |  |  |  |
| Gender (F/M) | χ² = 0.71 | | | |  | .398 |  | ns |
| Age (years) | 28.99 | 7.95 | 27.87 | 6.95 | 0.73 | .465 | +0.15 | ns |
| SOFAS | 60.29 | 12.52 | 55.42 | 15.90 | 1.65 | .101 | +0.36 | ns |
| PANSS positive | 6.00 | 3.13 | 6.05 | 3.19 | −0.08 | .935 | −0.02 | ns |
| PANSS negative | 6.24 | 3.34 | 5.90 | 3.44 | 0.51 | .610 | +0.10 | ns |
| TLI impoverishment | 0.31 | 0.36 | 0.47 | 0.54 | −1.67 | .098 | −0.37 | ns |
| TLI disorganisation | 0.46 | 0.39 | 0.60 | 0.45 | −1.57 | .120 | −0.34 | ns |
| LSM autobiog | 0.86 | 0.04 | 0.85 | 0.04 | 1.01 | .315 | +0.22 | ns |
| LSM external | 0.76 | 0.06 | 0.74 | 0.07 | 1.18 | .239 | +0.26 | ns |
| LSM overall | 0.85 | 0.03 | 0.84 | 0.04 | 1.34 | .183 | +0.29 | ns |

Comparison of patients retained at 12-month follow-up (n = 70) versus those lost to follow-up (n = 39) on baseline clinical and LSM variables. Independent-samples t-tests for continuous variables; chi-square for gender. d = Cohen's d; p = p-value. SOFAS = Social and Occupational Functioning Assessment Scale; PANSS = Positive and Negative Syndrome Scale; TLI = Thought and Language Index; LSM = linguistic style matching. ns = non-significant.

**Table S1. Attrition comparison: retained vs. lost-to-follow-up patients at baseline**
