## Supplementary material for "Two-Person Psychopathology: Linguistic Style Matching Marks Disorganized Self-Referential Narrative in Psychosis": Table S2

| **High LSM LSM = 0.925** | |
| --- | --- |
| **INTERVIEWER** | Can you tell me a story about something important that happened to you in your life? |
| **PARTICIPANT** | Something important. Wow. The most important thing that happened in my life was... the passing of my mother. There's a hole in my heart and I don't know how to fill it. It's been very difficult lately. |
| **INTERVIEWER** | Did you want to maybe share some of your favourite memories that relate back to your mom? |
| **PARTICIPANT** | Yeah, I remember when she bought me my first Super Nintendo. That really changed my life — even though things weren't going well at school, I could always turn to those games for happiness. It was a place where I could feel safe. |
| **INTERVIEWER** | It's definitely a happy memory, yeah. |

| **Low LSM LSM = 0.760** | |
| --- | --- |
| **INTERVIEWER** | So first, can you tell me a bit about yourself? |
| **PARTICIPANT** | Sure. I'm currently a student. I live, I suffer, and I breathe. That's kind of the reference — but yeah, I make mistakes, I have flaws, and I'm trying to fix it. |
| **INTERVIEWER** | Do you want to maybe talk a bit more about your studies? |
| **PARTICIPANT** | I'm doing graphic design. I want to express myself, get what's on my mind off of it — share what I have to say. |
| **INTERVIEWER** | And is that mostly on computer or actual art form on paper? |
| **PARTICIPANT** | It's on computer, but I'm trying to sketch it out first — if you just do it on the computer it'll look like a child's thing, and mine currently does, but I'm trying to make it look better. |

Verbatim excerpt illustrating linguistic style matching (LSM) between interviewer and participant. LSM quantifies unconscious function-word mirroring between speakers (Niederhoffer & Pennebaker, 2002). Scores range 0–1; higher = greater synchrony. Participant turns shaded for readability.

**Table S2. High/Low LSM verbatim excerpt**
