## Supplementary material for "Two-Person Psychopathology: Linguistic Style Matching Marks Disorganized Self-Referential Narrative in Psychosis": Table S3

| **Outcome** | **Predictor** | **β** | **SE** | **d** | **p** | **n** | **Sig.** |
| --- | --- | --- | --- | --- | --- | --- | --- |
| **Autobiog LSM** | **PT token count (autobiog)** | **0.0092** | **0.0029** | **0.23** | **.002** | 101 | **p < .05 *** |
| External LSM | PT token count (external) | 0.0040 | 0.0052 | 0.06 | .445 | 100 | ns |
| **Overall LSM** | **PT token count (autobiog)** | **0.0108** | **0.0027** | **0.29** | **<.001** | 101 | **p < .05 *** |

**.** Linear mixed-effects model (random intercept per participant) testing whether participant word output (token count, z-scored) predicts LSM independent of timepoint (tp_c centred at 0). Patients only. β = coefficient for token count z-score; d = Cohen's d effect size; n = number of participants. * p < .05; ns = non-significant.

**Table S3. Verbosity LMER: participant token count predicting LSM independent of timepoint**
