## Supplementary material for "Two-Person Psychopathology: Linguistic Style Matching Marks Disorganized Self-Referential Narrative in Psychosis": Table S5

| **Component** | **Eigenvalue** | **% Variance** | **Cumulative %** | **ΔPANSS Positive** | **ΔPANSS Negative** | **ΔSOFAS** | **ΔTLI Impoverishment** | **ΔTLI Disorganisation** |
| --- | --- | --- | --- | --- | --- | --- | --- | --- |
| **cPC1** | **1.774** | **34.8** | **34.8** | **+0.556** | **+0.497** | **+0.596** | +0.035 | -0.295 |
| **cPC2** | **1.151** | **22.6** | **57.4** | -0.172 | -0.024 | -0.146 | **+0.791** | **-0.568** |
| **cPC3** | **0.919** | **18.1** | **75.5** | -0.355 | **+0.542** | +0.162 | **+0.413** | **+0.619** |

Change-score Clinical PCA = 12m - baseline (n = 56 patients). Three components retained: two met the Kaiser criterion (eigenvalue > 1; bold rows); a third (eigenvalue = 0.919) was additionally retained on theoretical grounds given the interpretive coherence of a negative-disorganisation trajectory. KMO = 0.589; Bartlett's χ²(10) = 22.14, p = .014. SOFAS sign-inverted so that higher values indicate decline. Loadings extracted by PCA without rotation. Bolded cells indicate |loading| ≥ 0.40. cPC1 contrasts general symptom improvement (↓ PANSS, ↑ SOFAS) with stability; cPC2 primarily loads on TLI disorganisation change; cPC3 captures a negative-disorganisation trajectory (↑ PANSS negative, ↑ TLI disorganisation).

**Table S5. Change-score PCA**
