## Supplementary material for "Two-Person Psychopathology: Linguistic Style Matching Marks Disorganized Self-Referential Narrative in Psychosis": Table S6

| **Change-PC** | **Context** | **Pearson r** | **Pearson *p*** | **GEE β** | **95% CI** | **Cohen's d** | **GEE p** |
| --- | --- | --- | --- | --- | --- | --- | --- |
| cPC1 | Overall | +0.041 | .764 | −0.005 | −0.020, +0.010 | −0.14 | .533 |
| cPC1 | Autobiog | +0.046 | .739 | −0.005 | −0.018, +0.008 | −0.12 | .457 |
| cPC1 | External | +0.113 | .415 | +0.004 | −0.011, +0.020 | +0.07 | .569 |
| cPC2 | Overall | −0.030 | .826 | −0.002 | −0.013, +0.0095 | −0.06 | .716 |
| cPC2 | Autobiog | −0.018 | .898 | +0.002 | −0.012, +0.016 | +0.05 | .773 |
| cPC2 | External | −0.107 | .440 | −0.006 | −0.017, +0.003 | −0.10 | .218 |
| **cPC3** | **Overall** | **+0.437** | **<.001** | **+0.018** | **+0.005, +0.031** | **+0.50** | **.006** |
| **cPC3** | **Autobiog** | **+0.297** | **.028** | **+0.017** | **+0.001, +0.034** | **+0.43** | **.031** |
| cPC3 | External | +0.208 | .131 | +0.015 | −0.001, +0.032 | +0.24 | .079 |

**Note.** Pearson correlations between each change-PC score and RTM-corrected LSM change (ΔLSM_adj) were computed separately for each discourse context (overall/autobiographical n = 55; external n = 54). Generalized Estimating Equations (GEE; n = 56) with an exchangeable correlation structure and robust sandwich standard errors tested the corresponding longitudinal association using the model LSM ~ timepoint + PC_z + timepoint × PC_z. β = unstandardized coefficient for the timepoint × PC_z interaction term; 95% CI = confidence interval; d = Cohen's d; PC_z = standardized change-PC score. cPC1 = general improvement; cPC2 = disorganisation; cPC3 = negative-disorganisation trajectory.

**Table S6. Prospective associations between clinical change dimensions and longitudinal LSM**
