## Supplementary material for "Two-Person Psychopathology: Linguistic Style Matching Marks Disorganized Self-Referential Narrative in Psychosis": Table S7

| **Change-PC** | **r** | **p** | **n** | **sig.** |
| --- | --- | --- | --- | --- |
| **cPC1 (General improvement)** | **+0.333** | **.024** | 46 | ***** |
| cPC2 (Disorganisation) | +0.137 | .364 | 46 | ns |
| cPC3 (Neg-disorg. trajectory) | −0.193 | .199 | 46 | ns |

Pearson correlations between change-PC scores and ΔDDD (DDD_AP_12m − DDD_BL) in patients with complete PCA and antipsychotic dose data (n = 46 of 56 PCA-complete patients). ΔDDD descriptives: mean = −0.107, SD = 0.739, range = −2.500 to 1.625; escalated n = 19, reduced n = 25, unchanged n = 2. cPC1 = general improvement dimension; cPC2 = disorganisation dimension; cPC3 = negative-disorganisation trajectory. * p < .05; ns = non-significant.

**Table S7. Antipsychotic dose change (ΔDDD) × change-PC scores** — Pearson correlations
